# Integrated Early Childhood Development Centres in Market and Cross-Border Settings: a Mixed-Methods Evaluation in Rwanda

**DOI:** 10.64898/2026.05.14.26353227

**Authors:** Eric Matsiko, Pierre Nzeyimana, Annet Birungi, Samson Desie

**Author notes:** These authors contributed equally to this work (Eric Matsiko, Samson Desie). These authors contributed equally to this work (Pierre Nzeyimana, Annet Birungi).

## Abstract

**Introduction:** Access to quality early childhood development (ECD) services remains limited for families working in economic settings in many low-and middle-income countries. This study examined the associations between participation in integrated ECD centres of integrated ECD centres located in market and cross-border communities in Rwanda and childcare access, child nutrition, caregiving practices, and developmental outcomes.

**Methods:** A repeated cross-sectional pre–post evaluation without a comparison group was conducted between 2023 and 2025 across eight markets and cross-border ECD centres in Rwanda. Quantitative data were analyzed with logistic regression models adjusted for socio-demographic characteristics, while qualitative data were analysed thematically. University of Rwanda College of Medicine and Health Sciences Institution review board approved the study (No.366/CMHS IRB/2023).

**Results:** The proportion of children aged 6–23 months who achieved minimum meal frequency increased from 26.6% to 57% (AOR=2.35; 95% CI: 1.01–5.49), and those meeting minimum acceptable diet increased from 15.4% to 51.4% (AOR=4.51; 95% CI: 1.77–11.45). Stunting declined from 32.6% to 15.7% (AOR=0.45; 95% CI: 0.32–0.63) and underweight from 9.3% to 4.3% (AOR=0.55; 95% CI: 0.32–0.97). The proportion of children aged 24–59 months who were developmentally on track increased from 77.9% to 93.8% (AOR=3.85; 95% CI: 2.23–6.65). Households reported higher income at endline, and the centres generated strong demand for childcare services. However, reports of scolding and physical punishment increased between surveys.

**Conclusions:** Integrated ECD centres in market and cross-border settings were associated with improved child feeding practices, nutritional status, and developmental outcomes among children from vulnerable working families in Rwanda. Place-based childcare models may represent a promising strategy for expanding access to integrated ECD services while supporting women’s economic participation in economic settings.

## Introduction

The early childhood period is characterized by rapid physical, cognitive, and socio-emotional development, with long-lasting implications for health, educational attainment, and economic productivity across the life course (1, 2). However, millions of children in low- and middle-income countries remain at risk of poor developmental outcomes due to poverty, inadequate nutrition, limited opportunities for early learning, and exposure to unsafe or unstimulating caregiving environments(1). The Nurturing Care Framework emphasizes the importance of integrated interventions that address adequate nutrition, health, responsive caregiving, opportunities for early learning, and safety and security to support optimal child development (2, 3).

In response to these challenges, there has been increasing global recognition of the importance of integrated ECD services. Multisectoral ECD approaches combine early learning opportunities with nutrition support, health promotion, parenting education, and child protection services to address the multiple determinants of child wellbeing (2, 3). Evidence from low- and middle-income countries demonstrates that integrated ECD programmes can improve child nutrition, cognitive development, and school readiness, particularly among vulnerable populations (4, 5).

Despite growing attention to ECD, access to quality childcare and early learning services remains limited in many low- and middle-income countries, particularly for families engaged in economic activities (6, 7). Parents working in markets and cross-border trade often face long working hours, mobility constraints, and limited access to affordable childcare services (6, 8–10). In such contexts, young children may remain in unsafe or poorly supervised environments, limiting opportunities for adequate nutrition, stimulation, and responsive caregiving (1). Rapid urbanization and the expansion of informal economies across sub-Saharan Africa have further increased the demand for flexible and accessible childcare solutions for working families (6, 7).

In Rwanda, early childhood development has been prioritized within the national human capital development agenda. The Government of Rwanda has adopted a multisectoral ECD policy that promotes integrated services across health, nutrition, early learning, water and sanitation, and child protection sectors (11). To respond to diverse community needs, Rwanda has implemented multiple ECD service delivery models, including community-based, school-based, home-based, faith-based, work-based, market-based, and cross-border ECD centres (12). Within this framework, UNICEF and its partners have supported the establishment of ECD centres located within markets and cross-border trading areas to address childcare challenges faced by working parents, particularly women engaged in trade. These centres provide safe environments where children can access early learning, nutrition, health, and protection services while caregivers engage in economic activities.

Although evidence on community-based ECD interventions has expanded globally, empirical evidence on place-based childcare models embedded within economic environments remains limited, particularly in sub-Saharan Africa (3, 8). Relatively little is known about the potential contribution of ECD centres located directly within markets and cross-border trading settings to child nutrition, developmental outcomes, caregiving practices, and childcare access among vulnerable working families.

This study, therefore, examined the association between integrated ECD centres established in market and cross-border settings in Rwanda and child feeding practices, nutritional status, developmental outcomes, caregiving practices, and childcare access. By focusing on ECD services embedded in economic environments, the study contributes evidence on the potential role of place-based childcare models in improving child wellbeing while supporting women’s economic participation in low-resource settings.

## Materials and Methods

### Study Design

The study used a repeated cross-sectional pre–post evaluation without a comparison group design. At the endline, qualitative data using focus group discussions (FGDs), in-depth interviews (IDIs), and key informant interviews (KIIs) were collected. The endline assessment was conducted in August 2025 and followed a baseline survey carried out in October 2023, before the implementation of project activities.

### Study setting

The market-based sites were Musanze Market (Musanze District), Gihango Market (Rutsiro District), Gakenke Market (Gakenke District), Kimironko Market (Gasabo District), and Zinia Market (Kicukiro District). The cross-border sites included Rusizi (Rusizi District), Kagitumba (Nyagatare District), and Gatuna (Gicumbi District).

### Study Participant

Eligible participants included children aged 6 months to 6 years and their caregivers (parents or guardians), as well as ECD caregivers, community health workers (CHWs), and other key stakeholders involved in ECD service delivery.

### Sampling and sample size

At endline, lists of children enrolled in the ECD centres were obtained from the implementing partner and used to identify eligible child–caregiver pairs. All children attending the centres at the time of fieldwork were included in the endline survey. At baseline, a sampling frame of potential project participants was developed with the support of community health workers (CHWs), from which a random sample of eligible child–caregiver pairs was selected

The sample size was calculated using a single-proportion formula adjusted for design effect, assuming a national stunting prevalence of 33% among children under five in Rwanda in 2020 (13), a 5% margin of error, a 95% confidence level, and a design effect of 1.5. A 10% non-response allowance was added to account for refusals, respondent unavailability, incomplete interviews, and missing or invalid anthropometric measurements. Based on these assumptions, the required sample size was 566 children. The target sample size was achieved at baseline. However, at endline, the intended sample size could not be fully reached due to a lower-than-anticipated number of children enrolled in the project ECD centres at the time of fieldwork. As a result, data were collected from 422 child–caregiver pairs at endline.

The qualitative component employed purposive sampling. One FGD was conducted at each project site with parents or guardians, resulting in a total of eight FGDs. IDIs were conducted with parents or guardians, community health workers, and ECD caregivers at each site. The number of FGDs and IDIs conducted was guided by the principle of data saturation, whereby interviews continued until no substantial new themes emerged. Key informant interviews included district ECD focal persons, national-level ECD stakeholders ((National Child Development Agency (NCDA), UNICEF)), and representatives of the project implementing partner. Structured observations were also conducted in all eight ECD centres.

### Data collection

The study employed a mixed-methods approach combining quantitative and qualitative data collection. Quantitative data were collected through structured household questionnaires administered to parents or guardians of children aged 6 months to 6 years. Anthropometric measurements were taken by trained enumerators following WHO standard procedures, and child development was assessed using the Early Childhood Development Index 2030 (ECDI2030) (14). Feeding practices among children aged 6–23 months were assessed according to the 2021 World Health Organization (WHO) guidelines (15). Data were collected electronically using the KoboCollect application, with daily checks conducted to ensure completeness and accuracy.

FGDs were conducted with approximately eight parents or guardians at each ECD centre. The discussions were held within the ECD centre facilities and were facilitated by an experienced moderator, assisted by a note-taker. The same research team conducted IDIs with one parent or guardian, one community health worker, and one ECD caregiver at each study site. In addition, the team carried out structured observations at each ECD centre to assess infrastructure, WASH conditions, nutrition and health services, the learning environment, staffing, and safety conditions. Qualitative data from FGDs, IDIs, and KIIs were audio-recorded with participants’ consent.

All data collectors received standardized training on the study objectives, data collection tools, ethical procedures, and child safeguarding before fieldwork.

### Outcome measures

Nutritional status indicators—height-for-age (HAZ), weight-for-height (WHZ), and weight-for-age (WAZ), were calculated using the WHO Anthro software, which applies the 2006 WHO Child Growth Standards(16). Stunting, underweight and wasting were defined as HAZ, WAZ and WHZ values below −2 standard deviations (SD) from the WHO reference population median, respectively. Complementary feeding indicators, including minimum meal frequency (MMF), minimum dietary diversity (MDD), and minimum acceptable diet (MAD), were computed in accordance with WHO guidelines (15). Child development outcomes were assessed using the Early Childhood Development Index 2030 (ECDI2030) framework (14). Overall, the outcome variables were selected in alignment with the Nurturing Care Framework, covering key domains of adequate nutrition, health, early learning, responsive caregiving, and safety and security.

### Data analysis

Quantitative data were cleaned and analyzed using Stata software. Descriptive statistics summarized the characteristics of the study population, including caregiver demographics and household characteristics. Continuous variables were presented using medians and interquartile ranges (IQR), while categorical variables were summarized as frequencies and percentages. To assess associations between programme implementation and child outcomes, logistic regression models were fitted for binary outcome variables. Survey round (baseline vs endline) served as a proxy for programme exposure, allowing examination of changes in key outcomes following the implementation of ECD services. Separate logistic regression models were estimated for each binary outcome variable and were adjusted for socio-demographic and household characteristics, including caregiver age, caregiver literacy status, household headship, and household income. Multicollinearity among independent variables was assessed using variance inflation factors (VIF), with all values below 2 indicating no evidence of multicollinearity. Robust standard errors were used to account for potential heteroskedasticity, and statistical significance was assessed at the 5% level. Adjusted odds ratios (AORs) with 95% confidence intervals were reported to estimate the magnitude and direction of associations. The analysis aimed to assess associations between exposure to the ECD programme and key child and household outcomes over time rather than to establish causal effects.

Missing data were minimal and were addressed through complete-case analysis, whereby only records with available information for the variables of interest were included in the final analysis. Data cleaning and consistency checks were conducted prior to analysis to minimize errors and incomplete responses. No imputation methods were applied because the proportion of missing data was low (< 1%).

Recorded qualitative data were transcribed verbatim and reviewed against the original recordings for accuracy. Analysis followed an inductive thematic approach. To enhance reliability, two authors independently coded a subset of transcripts and harmonized the coding framework through discussion. The agreed coding framework was then applied to the remaining transcripts. Emerging themes were refined and organized into overarching categories, and illustrative quotations were used to highlight key findings and provide contextual depth.

### Ethics

The study followed established ethical standards for research involving human participants, including the Declaration of Helsinki and child safeguarding guidelines. The University of Rwanda Institutional Review Board approved the study protocol (No. 366/CMHS IRB/2023) before data collection. Written informed consent was obtained after researchers informed participants about the study objectives, procedures, risks, and benefits and obtained informed consent before participation. Participation was voluntary, and participants could withdraw at any time without consequences. The research team ensured confidentiality by removing personal identifiers, restricting access to raw data, and storing electronic data securely in password-protected systems.

## Results

### Participant characteristics

**Table 1** displays the socio-demographic and economic characteristics of parents/guardians and their households at both baseline and endline. The proportion of parents/guardians who were mothers was higher at endline (89%) than at baseline (83.7%) (p = 0.017). The median age of caregivers was also greater at endline, at 32 years (IQR: 9.5), compared to 30 years (IQR: 9) at baseline (p = 0.015). Caregiver literacy was higher at endline (91.2%) than at baseline (87%) (p = 0.038). Household composition varied between survey rounds, with male-headed households constituting a larger share at endline (78%) compared to baseline (67%) (p = 0.000). Household economic features also changed between surveys. Household economic characteristics differed between surveys, with a larger proportion of households reporting higher monthly income at endline (50.5%) compared to baseline (20%) (p = 0.000). Conversely, the distribution of marital status, household size, and the number of young children remained similar across both baseline and endline surveys.

**Table 1:**
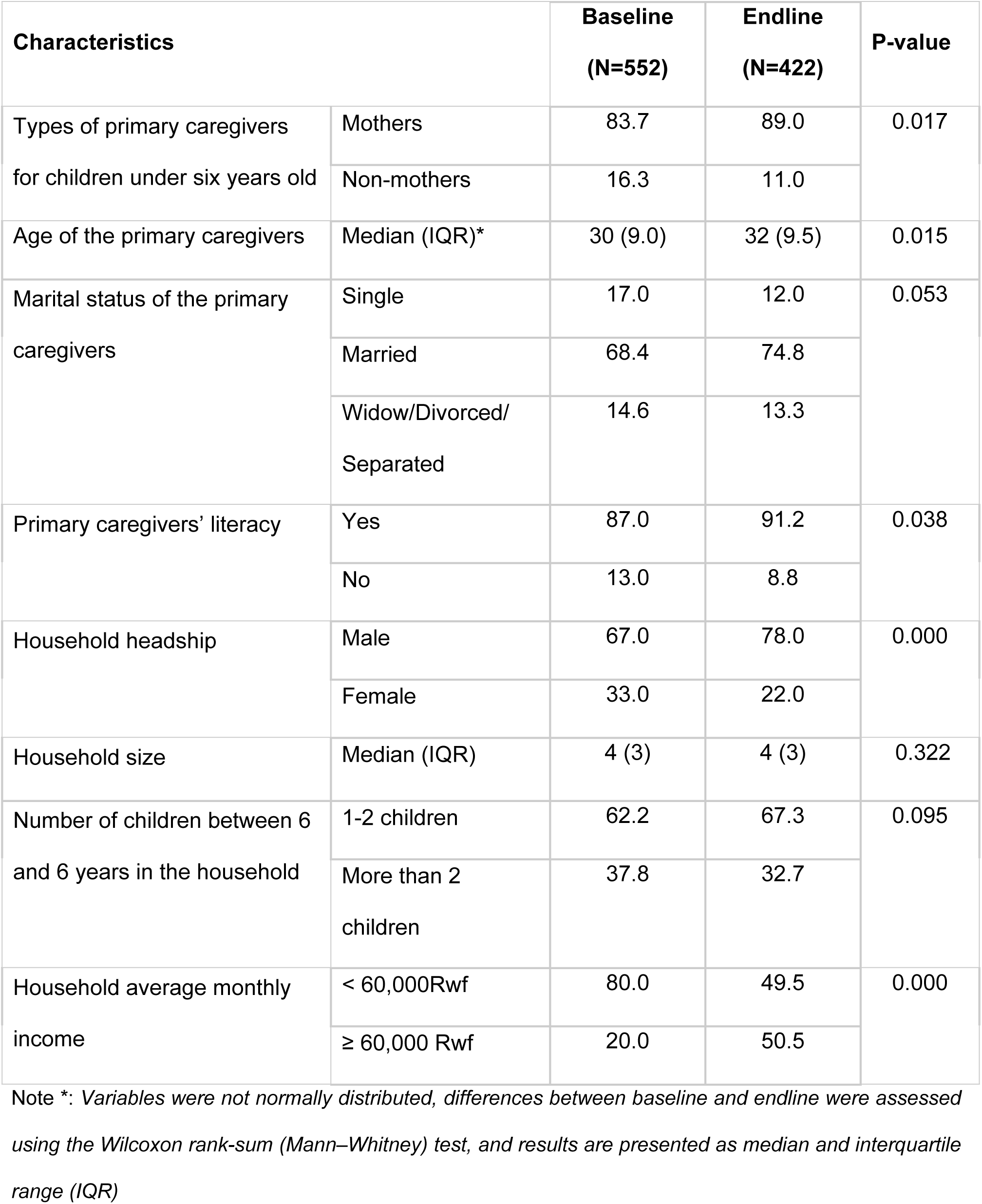
Socio-demographic and economic characteristics of the participants (primary caregivers) and their households.

### Access to and quality of integrated ECD services

The project expanded access to ECD services in underserved markets and cross-border areas by establishing eight operational centres: five market-based (Kimironko, Gakenke, Musanze, Congo-Nil/Rutsiro, and Ziniya) and three cross-border (Kagitumba, Gatuna, and Rusizi). Over two years, the centres provided safe childcare and integrated ECD services to 701 children, exceeding the target of 640. Strong community demand in all locations often surpassed centre capacity, indicating a need for further expansion.

Overall service quality improved and generally met national ECD minimum standards. Centres delivered integrated services including early learning, nutrition, health, hygiene, child protection, and positive parenting (17). Most had child-friendly environments with sanitation facilities, cooking and feeding areas, and learning materials. Safety conditions varied across sites, with some centres demonstrating strong standards (adequate space, ventilation, cleanliness, secure fencing), while others faced minor gaps such as overcrowding, weak fencing, uneven play areas, poor ventilation, and inadequate waste management. Collaboration with community health workers supported growth monitoring, immunization follow-up, hygiene promotion, and nutrition education. However, the absence of first aid kits and formal referral protocols limited preparedness for medical emergencies.

Human resource capacity improved through training in inclusive play, nurturing care, disability-responsive services, and early learning facilitation. Despite these gains, service quality was constrained by high child-to-caregiver ratios and caregiver shortages linked to low or irregular incentives. Caregivers implemented structured daily routines that supported early learning, stimulation, rest, and feeding, contributing to improved developmental outcomes. However, infrastructure limitations in some centres continued to affect optimal play and learning conditions.

### Child nutrition and development outcomes

Changes in feeding practices, nutritional status, and developmental outcomes among children are presented in **Table 2**. The proportion of children aged 6–23 months meeting minimum meal frequency was higher at endline (57%) than at baseline (26.6%), and adjusted analyses indicated higher odds of meeting minimum meal frequency at endline (AOR = 2.35; 95% CI: 1.01–5.49). Similarly, the proportion of children meeting minimum acceptable diet criteria was higher at endline (51.4%) than at baseline (15.4%), with adjusted analyses indicating significantly higher odds at endline (AOR = 4.51; 95% CI: 1.77–11.45). Although minimum dietary diversity among children aged 6–23 months was higher at endline (66.7%) than at baseline (44%), adjusted analyses indicated no significant difference.

**Table 2:** Child nutrition and development outcomes.

| Characteristics | Baseline<br>(n, %) | Endline<br>(n, %) | AOR (95% CI) * | P-value |
| --- | --- | --- | --- | --- |
| **Contribution to adequate nutrition** |  |  |  |  |
| Minimum Meal Frequency (6-23 months) | 51 (26.6) | 20 (57.0) | 2.35 (1.01-5.49) | 0.041 |
| Minimum Dietary Diversity (6-23 months) | 92 (44.0) | 24 (66.7) | 2.10 (0.90-4.92) | 0.086 |
| Minimum Acceptable Diet (6-23 months) | 30 (15.4) | 18 (51.4) | 4.51 (1.77-11.45) | 0.001 |
| Animal-source food consumption among children aged 6-72 months | 281 (52.8) | 258 (47.2) | 1.16 (0.86-1.55) | 0.312 |
| **Contribution to nutrition status and child development** |  |  |  |  |
| Stunting | 179 (32.6) | 66 (15.7) | 0.45 (0.32–0.63) | 0.000 |
| Underweight | 51 (9.3) | 19 (4.5) | 0.55 (0.32-0.97) | 0.039 |
| Wasting | 11 (2.2) | 17 (4.0) | 1.94 (0.84-4.51) | 0.120 |
| Developmentally on track among children aged 24-59 months | 240 (77.9) | 322 (93.8) | 3.85 (2.23-6.65) | 0.000 |

**Note:** \*\*Each individual model was adjusted for the type of primary caregivers, Age of the primary caregivers, the primary caregivers' literacy, Household Headship and household Income. AOR: Adjusted Odds Ratio. CI: Confidence Interval.

With respect to nutritional status and child development (**Table 2**), the prevalence of stunting was lower at endline (15.7%) compared with baseline (32.6%), with adjusted analyses indicating lower odds of stunting among children assessed at endline (AOR = 0.45; 95% CI: 0.32–0.63). Similarly, prevalence of underweight at endline was lower compared with baseline (4.3% vs. 9.3%), with adjusted analysis showing significantly lower odds of being underweight at endline (AOR = 0.55; 95% CI: 0.32–0.97). In terms of child development, the proportion of children aged 24–59 months who were classified as developmentally on track increased from 77.9% at baseline to 93.8% at endline. Adjusted regression results further indicated significantly higher odds of being developmentally on track among children assessed at endline (AOR = 3.85; 95% CI: 2.23–6.65).

In FGDs and interviews, parents consistently highlighted nutrition and developmental benefits of the ECD centres, which motivate them to remain engaged and actively support the centres. Parents explained that meals at home were often irregular, whereas children at the centres receive more balanced diets including vegetables, fruits, eggs, and porridge. Caregivers also reported noticeable improvements in children’s growth and vitality. As one caregiver explained, “*The changes are clear: children who were behind in growth indicators improved*. *They became stronger, more energetic, and happy to come to the centre”* (Caregiver, Gakenke). A parent similarly noted tangible improvements in weight gain: *“My child was not gaining weight, but since joining the centre, he has increased from 9 kg to 13 kg. The food offered at the center helped him a lot”* (Parent, Gicumbi). These observations are consistent with the quantitative findings, which show substantial reductions in child underweight between baseline and endline.

Parents and caregivers also emphasized significant progress in children’s learning and communication skills. Children were reported to gain abilities in speaking, counting, singing, and learning basic words in different languages. As one parent shared*, “My child came here without speaking, but now he sings songs, counts in English and Kinyarwanda, and even asks for things clearly. This gives me confidence for his future”* (Parent, Rusizi cross-border market). Another parent noted improvements in confidence and interaction: *“The most helpful thing is that he gained confidence, interacts with other children, and has learned to speak and count”* (Parent, Kimironko market). These qualitative experiences align with the quantitative results showing a marked increase in the proportion of children aged 24–59 months who were developmentally on track at endline.

Caregivers attributed these gains to play-based learning approaches used at the centres, including songs, poems, and counting games. As one caregiver explained, *“We use songs, poems, and counting games so children learn while playing”* (Caregiver, Gatuna cross-border market).

Beyond learning, the centres were also associated with children’s social development. Parents and caregivers observed that children became more confident, expressive, and socially engaged. One parent explained, *“My child used to be shy and withdrawn, but since joining the ECD, he plays freely, communicates his needs, and even tells me what the teacher taught him*” (Parent, Gakenke). Another parent highlighted changes in behaviour at home: *“My child never played, but now he plays football, sings, and even reminds us to wash our hands before eating”* (Parent, Gakenke market). Together, these experiences complement the quantitative evidence, illustrating how the centres contribute to improvements in children’s nutrition, development, and social well-being while strengthening parents’ confidence in their children’s future.

#### Health and Hygiene Practices and attending parenting sessions

**Table 3** presents differences in health and hygiene-related practices and participation in parenting activities between baseline and endline surveys. Among health indicators, coverage of deworming remained high and comparable across survey rounds (91% at baseline and 94.7% at endline). Care-seeking during child illness was also similar, increasing slightly from 83.8% at baseline to 88.6% at endline. In contrast, vitamin A supplementation declined from 95.3% at baseline to 89% at endline, with adjusted analyses indicating lower odds at endline (AOR = 0.35; 95% CI: 0.21–0.58). The proportion of children reported to have experienced diarrhea in the two weeks preceding the surveys also declined from 33% at baseline to 25.3% at endline, but this decline was not significant between surveys.

**Table 3:**
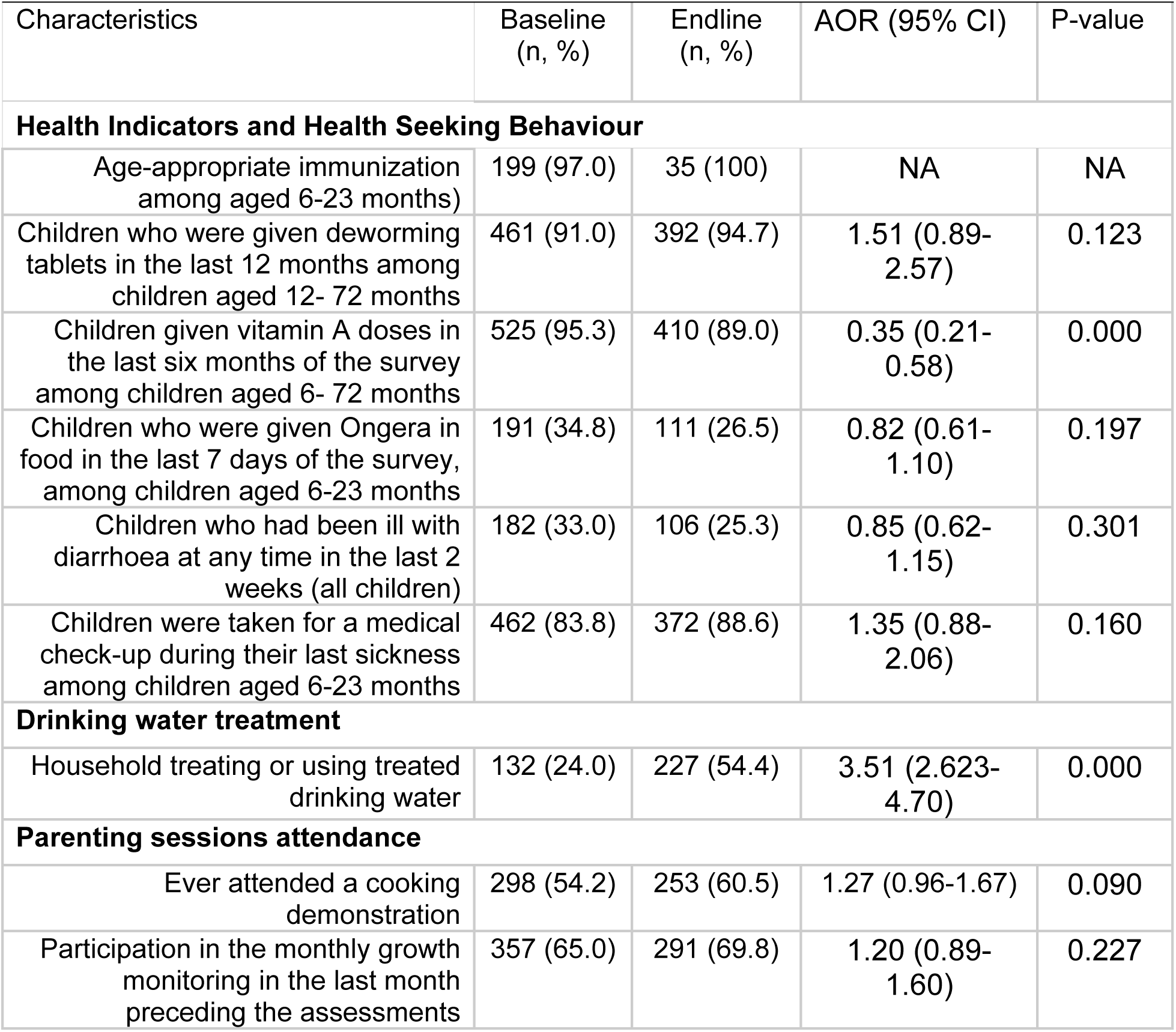
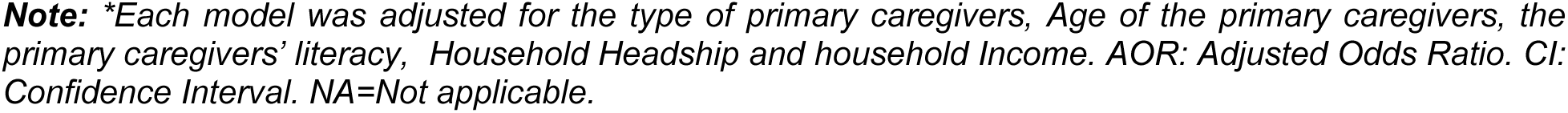
Health and Health-Seeking Behaviour, water treatment and Parenting Sessions.

Substantial improvements were observed in household drinking water treatment practices. The proportion of households treating or using treated drinking water increased significantly from 24% at baseline to 54.4% at endline (AOR = 3.53; 95% CI: 2.62–4.70). Additionally, qualitative insights showed that hygiene practices have improved. Children were then familiar with washing their hands before eating and maintaining cleanliness throughout the day. Parents observed tangible spillover effects at home, as children now remind their siblings and even adults about handwashing. A parent in Gakenke market noted, *“Even at home, my child refuses food unless he washes his hands first. This has also taught me to always wash my hands before giving him food.”* Similarly, parents in Kagitumba cross-border market highlighted the contrast in children’s appearance, with one remarking, *“Children here are bathed and dressed in clean clothes. When you come to pick them up, you find them neat, unlike before when they roamed the village dirty.”*

Participation in parenting-related activities showed modest changes between survey rounds (Table 3). Attendance at cooking demonstrations increased slightly from 54.2% at baseline to 60.5% at endline, while participation in monthly growth monitoring rose from 65% to 69.8%. However, these differences were not statistically significant. Qualitative findings suggest that limited parental participation may reflect the demanding work schedules of market and cross-border traders. As one community health worker explained, “*Teaching parents here is difficult because many are busy, leaving their children early in the morning and returning in the evening”* (CHW, Gatuna Cross-border).

#### Child protection outcomes

**Table 4** presents differences in child protection and safety practices between the baseline and endline surveys. Two child discipline indicators showed significant differences between the survey rounds. The proportion of children reported to be scolded at home increased markedly from 34.8% at baseline to 78.5% at endline, with adjusted analyses showing substantially higher odds at endline (AOR = 9.27; 95% CI: 6.54–13.11). Similarly, the proportion of children reported to have experienced physical punishment increased from 61.6% at baseline to 68% at endline, and adjusted estimates likewise indicated significantly higher odds at endline (AOR = 1.43; 95% CI: 1.07–1.90). For the remaining indicators, including children being insulted at home, left alone at home, left in the care of another child younger than 10 years, or left in the care of a non-related household member, differences observed between baseline and endline were not statistically significant.

**Table 4:** Child protection and safety indicators.

| Characteristics | Baseline<br>(n, %) | Endline<br>(n, %) | AOR (95% CI) | P-value |
| --- | --- | --- | --- | --- |
| Children being scolded at home | 191 (34.8) | 382 (78.5) | 9.27 (6.54-13.11) | 0.000 |
| Children being physically punished | 338 (61.6) | 285 (67.8) | 1.43 (1.07-1.90) | 0.013 |
| Children being insulted at home | 306 (56.2) | 200 (47.6) | 0.88 (0.66-1.16) | 0.387 |
| Children were left alone at home | 172 (31.3) | 116 (27.6) | 0.95 (0.69-1.30) | 0.762 |
| Children being left in the care of another child less than 10 years old | 230 (41.7) | 173 (41.2) | 1.06 (0.80-1.40) | 0.664 |
| Children being left in the care of another household member who is not related to them | 296 (53.7) | 217 (51.7) | 1.00 (0.76-1.32) | 0.972 |

**Note:** *\*Each model was adjusted for the type of primary caregivers, Age of the primary caregivers, the primary caregivers' literacy, Household Headship and household Income. AOR: Adjusted Odds Ratio. CI: Confidence Interval.*

Qualitative evidence indicates that parents strongly value the protective environment provided by the ECD centres. Parents emphasized the peace of mind they now experience compared with the past, when children were left with neighbors, house helpers, or allowed to roam in markets or fields. As one parent explained, “*Before, I left my child with neighbors and found them beaten or wandering. Now I leave them here safely and go to work”* (Parent, Kagitumba cross-border market). Another parent similarly noted, “*At the ECD, I feel at peace because I know he is safe, has eaten, and rested”* (Parent, Gatuna cross-border market).

The centres have also reduced risks in crowded markets and community spaces where children were previously unsupervised. One parent described how the centres now serve as safe points for children: “*At the market, children used to roam and get lost. Now, if children are seen wandering, people bring them to the ECD, and parents can find them quickly”* (Parent, Musanze market). Caregivers confirmed that children are continuously supervised in safe environments: “*A child here is protected; there are no dangerous objects, and caregivers are always present to ensure safety”* (ECD Caregiver, Musanze market).

Protection also extends to children’s emotional well-being. ECD Caregivers reported using positive discipline approaches rather than harsh punishment. As one caregiver explained, “*We apply positive parenting and do not allow children to be insulted or hit. Instead, we guide them calmly to protect their emotions*” (ECD Caregiver, Musanze market).

#### Systems and governance outcomes

The centres function as integrated platforms for delivering early learning, health, nutrition, WASH, and child protection services. Insights from FGDs and KIIs indicate that this coordination is most evident through collaboration with CHWs and local health facilities. CHWs regularly support growth monitoring, malnutrition screening, immunization follow-up, hygiene promotion, and nutrition education at the centres, ensuring that children access essential health and nutrition services through existing systems rather than parallel structures. Caregivers often multitask by teaching, feeding, and managing hygiene, while parents contribute by fetching water or providing supplies, creating a collaborative and efficient approach. However, this multitasking may risk overstretching ECD centres’ caregivers, particularly where staffing levels are limited. As one caregiver noted, *“Only one caregiver manages many children, which reduces service quality and efficiency”* (Caregiver, Gatuna cross-border market)

The centres also serve as referral entry points to nearby health facilities, strengthening linkages between the ECD, health, and nutrition sectors. Collaboration further extends to the agriculture sector through the establishment of kitchen gardens at the centres supported by local authorities and communities. These gardens contribute to children’s meals while helping to reduce operational costs.

Local government ownership is reflected in district-level leadership, facilitation, and oversight of the ECD centres. District authorities support the centres by identifying and providing facilities, supervising service delivery, and integrating them into district ECD coordination mechanisms. As a result, the centres are linked to Rwanda’s national ECD system and benefit from government training, supervision, and policy guidance similar to other ECD centres. Parents further appreciated the transparency and accountability of the centres, noting that they and local leaders can visit at any time to check on children’s well-being. As one caregiver noted, “Parents are free to come at any hour to see their children, and local leaders also check on hygiene and safety” (ECD Caregiver, Kimironko market).

Local governments also mobilize partners and communities to support center operations, including advocating for infrastructure improvements and allocating land for community gardens. However, ownership and support remain uneven across districts. While some districts have been proactive in supporting infrastructure upgrades and operational needs, delays in providing or renovating government-owned buildings in others highlight differences in local government responsiveness.

The evaluation further indicates that market-based and cross-border ECD centres were increasingly being integrated into district development plans and the national ECD system. Districts have incorporated these centres into their ECD planning frameworks, and coordination with the NCDA has strengthened national oversight and alignment with policy standards. Joint planning, supervision, and monitoring between local authorities and implementing partners have also helped reduce duplication and improve implementation efficiency.

However, integration into district budgets remained partial. Although some districts have begun allocating resources or providing in-kind support such as facilities, land, or utilities, the centres continue to rely heavily on external partners and NGOs for key inputs, including caregiver training, food supplies, and infrastructure improvements. According to a key informant, this reliance on external support, combined with limited district budget allocations, may constrain the long-term sustainability and scale-up of the centres. As one district focal person observed, *“Parents contribute little by little, but the teaching materials and running costs remain far higher than what they can sustain.”* If donor priorities shift or government budgets tighten, centres may struggle to maintain service quality.

#### Equity and Inclusion

The project demonstrates clear gender relevance, particularly in relation to women’s economic participation and caregiving responsibilities. Women dominate vending and cross-border trade and serve as the primary caretakers in most households. At endline, women accounted for over 96% of primary caregivers in the project group, highlighting the highly gendered nature of childcare responsibilities. The distribution of enrolled children was relatively balanced by sex but slightly skewed toward girls, who represented 52.6% of children at endline compared with 47.4% boys.

By situating ECD centres within markets and border posts, the project addressed a key constraint that previously limited women’s mobility and productivity: access to childcare. The project targeted economically vulnerable households engaged in market and cross-border trade, often characterized by unstable incomes and limited access to affordable childcare. At baseline, approximately 80% of households earned less than 60,000 RWF (about USD 40) per month, confirming appropriate targeting. Over the implementation period, economic outcomes showed substantial improvement, with average household monthly income differing significantly between the baseline and endline surveys (p < 0.001). Parents consistently linked these improvements to the availability of ECD services, which freed time for productive work and reduced reliance on costly private childcare or informal arrangements.

Affordability emerged as a key equity feature. Monthly ECD fees (approximately 10,000–12,000 RWF, or USD 5–8) were substantially lower than private childcare alternatives, which parents reported could cost 100,000–200,000 RWF (USD 70–140) per month. This improved access for low-income families; however, even modest fees remained challenging for the most economically vulnerable households. Community support mechanisms, including parental in-kind contributions, kitchen gardens, and shared centre maintenance, helped offset costs and strengthen local ownership. At the same time, reliance on parental contributions may risk excluding the poorest households without additional public or partner support.

Regarding disability inclusion, the project made initial efforts through ECD caregiver training and awareness-raising. ECD Caregivers received training on inclusive play and disability-responsive care, and centres were expected to accommodate children with diverse developmental needs. Across observed centres, inclusion of children with disabilities was generally reported, with many centres enrolling children with speech and communication difficulties. However, caregivers noted limited capacity to support more complex needs, and one centre reported referring children with disabilities to other facilities rather than enrolling them. While centres appeared generally open to inclusion, practical constraints remained. Gaps in specialized skills, adaptive learning materials, and inclusive infrastructure limited the extent to which inclusion could be fully realized. Overall, although disability inclusion was recognized in the project design and caregiver training, children with disabilities remain insufficiently supported.

## Discussion

This study examined the association of integrated ECD centres located in market and cross-border settings with childcare access, child nutrition, health practices, and developmental outcomes. Overall, the findings suggest that the intervention expanded access to childcare services for vulnerable households and was associated with improvements in child nutritional status, complementary feeding practices, developmental outcomes, and household economic conditions. However, challenges related to service quality, child protection practices, and sustainability remain.

One of the major findings was the strong demand for ECD services within the market and cross-border communities. The centres served more children than initially targeted, indicating substantial unmet need for accessible childcare services in these settings. This aligns with evidence showing that access to affordable childcare remains limited for families working in informal economic sectors in many low- and middle-income countries (1, 6, 7). Locating ECD centres directly within trading environments appears to reduce structural barriers to childcare access among working parents, particularly mothers engaged in market and cross-border trade. Improvements observed in household income may partly reflect the availability of reliable childcare services, which enabled caregivers to spend more time on income-generating activities while ensuring safe care for their children. Similar findings have been reported in studies linking affordable childcare access to women’s labour participation and economic productivity (7, 18).

The centres also functioned as integrated service platforms linking early learning, nutrition, hygiene promotion, and health monitoring. Such multisectoral approaches are increasingly recognized as essential for improving child wellbeing because ECD is shaped by interconnected determinants, including nutrition, health, stimulation, and caregiving environments(1, 2). Collaboration between ECD centres, community health workers, and local authorities reflects Rwanda’s national strategy of integrating ECD services within existing community and health systems (11, 19). Nevertheless, some centres experienced overcrowding and high child-to-caregiver ratios, which may compromise service quality. Previous studies similarly highlight caregiver training, supervision, and adequate staffing as important determinants of ECD programme effectiveness (8, 20).

The study documented improvements in minimum meal frequency and minimum acceptable diet among children aged 6–23 months. These gains may reflect the combined effects of structured feeding routines at the centres, nutrition education, and improved caregiver awareness of recommended feeding practices. Previous evidence demonstrates that integrated nutrition and behaviour change interventions can improve complementary feeding practices in low-resource settings (21–23). Improvements in child nutritional status observed in this study may similarly reflect the combined influence of regular meals, growth monitoring, health referrals, and improved caregiving practices provided through the ECD centres (1, 24). However, because the study used a repeated cross-sectional pre–post design without a control group, these findings should be interpreted as associations rather than causal effects.

Consistent with improvements in nutrition and caregiving environments, the study also documented substantial gains in child developmental outcomes. A higher proportion of children aged 24–59 months were developmentally on track at endline than at baseline. These improvements may reflect the contribution of structured early learning activities, play-based stimulation, and nurturing care provided within the centres. Daily routines involving songs, games, social interaction, and guided learning are widely recognized as important components of effective ECD programmes (2). Similar evidence from global ECD interventions shows that combining early stimulation with responsive caregiving can significantly improve developmental outcomes among young children in disadvantaged settings (4, 5).

The increase in reported scolding and physical punishment between baseline and endline should be interpreted cautiously. One possible explanation is increased caregiver awareness and willingness to disclose disciplinary practices following exposure to parenting and child protection messaging. Previous studies have shown that parenting interventions may initially increase reporting of harsh disciplinary behaviours before behavioural changes occur (25–27). At the same time, the findings may also suggest that parenting support interventions within the project were insufficient to reduce harsh discipline practices at the household level. These results, therefore, highlight the continued need to strengthen positive parenting interventions and caregiver support within ECD programmes.

The findings also highlight both progress and ongoing challenges related to sustainability. Although local government authorities demonstrated ownership through coordination and infrastructure support, many centres continued to rely heavily on external partners for operational resources, including caregiver training, food provision, and infrastructure improvements. Global evidence suggests that sustainable ECD programmes require strong government leadership, stable financing, and integration into national systems (2, 3, 28). Strengthening integration of market- and community-based ECD models into district budgets and national financing frameworks will therefore be important for long-term sustainability and scale-up.

The findings can further be interpreted through the Nurturing Care Framework, which emphasizes adequate nutrition, good health, responsive caregiving, early learning opportunities, and safety and security as essential components of optimal child development (2). The integrated ECD centres addressed these domains simultaneously by providing meals, health monitoring, structured learning activities, and safe caregiving environments. By focusing on childcare services embedded within market and cross-border economic environments, this study contributes evidence on place-based ECD models designed to support vulnerable working families engaged in informal economic activities.

This study has some limitations. The repeated cross-sectional pre–post design without a comparison group limits causal attribution of observed changes to the intervention. Differences between baseline and endline samples may also partly reflect variations in participant characteristics rather than programme effects. In addition, several indicators relied on caregiver self-report and may therefore be subject to recall and social desirability bias. The findings may not be generalizable beyond the market and cross-border trading communities. Finally, the study did not assess the quality of caregiver–child interactions or learning processes in detail, which are important dimensions of ECD quality.

## Conclusion

In conclusion, the findings suggest that integrated ECD centres located in market and cross-border environments can be associated with improvements in childcare access, child feeding practices, developmental outcomes, and household economic status. However, sustaining these benefits will require continued investments in caregiver training, infrastructure quality, positive parenting interventions, and government financing mechanisms. Expanding place-based childcare models within economic settings may represent a promising strategy for improving early childhood development outcomes while simultaneously supporting women’s economic participation in Rwanda and similar contexts.

## Data Availability

Data will be provided with minimal request

## Acknowledgements

The authors thank the Bainum Family Foundation for financial support for this study. We are also grateful to the Government of Rwanda, particularly the National Child Development Agency (NCDA) and local government authorities, for their collaboration and insights on ECD implementation. We further acknowledge the community participants and data collectors whose contributions made this study possible.

